# Normative intracranial EEG highlights epileptic abnormalities across wakefulness and sleep

**DOI:** 10.1101/2025.09.10.25335533

**Authors:** Akash R. Pattnaik, Ian Z. Ong, John Bernabei, Carlos A. Aguila, Alfredo Lucas, Daniel J. Zhou, Ryan Gallagher, William K. S. Ojemann, Mariam Josyula, Nina Petillo, Joel S. Stein, Kathryn A. Davis, Nishant Sinha, Erin C. Conrad, Brian Litt

**Author notes:** Correspondence to: Akash Ranjan Pattnaik.

## Abstract

There is great interest in using quantitative methods to localize epileptic networks from intracranial EEG, a vital part of care for patients with drug resistant epilepsy (DRE). In particular, there is evidence that using interictal data for this purpose, which could eliminate the need to record seizures, has great potential to reduce morbidity from precipitated seizures and to decrease length of stay. How much of this data is required for this purpose, and from what state(s) of consciousness, is not known. In this study we analyzed interictal intracranial EEG (iEEG) data from 30 subjects and compared it against normative reference iEEG derived from 106 additional patients. We summarized brain activity and connectivity by computing spectral power and coherence in 6 frequency bands and computed *z*-scores relative to normal features within the same anatomical region. We used a validated algorithm to estimate the sleep or wake state. We applied a cross-validated random forest model to assign predicted abnormality value to each channel for each state of wakefulness. To determine the effectiveness of this approach for each unseen patient, we computed the area under the precision recall curve (AUPRC) between predicted abnormality within and outside of the resection zone. We further identified associations between predicted abnormalities and neuropsychological testing performance, highlighting applications of quantitative biomarkers to epilepsy comorbidities. We found that subjects with good seizure outcome (Engel 1) at 2 years had higher AUPRC than subjects with poor seizure outcome for predicted abnormalities in N2 sleep (Mann-Whitney test; *p*_Holm=Bonferroni_ < 0.05). Combining features from wakefulness and NREM sleep best separated good and poor seizure outcome subgroups (Mann-Whitney test; *p*_Holm=Bonferroni_ < 0.05, Cohen’s *d* = 1.62). Combinations of wake and sleep abnormalities and interictal spikes explained the variance in pre-surgical neuropsychological testing (*R*^2^ = 0.57-0.58).

## Introduction

Epilepsy affects over 50 million people in the world, with about 20-30% resistant to treatment with antiseizure medications^1,2^. Surgery remains the gold standard therapy for drug-resistant epilepsy (DRE), and postoperative seizure freedom crucially relies on accurately identifying the network region responsible for generating seizures (epileptogenic zone (EZ))^3^. Surgical planning strategies rely on multimodal data including seizure semiology, electroencephalography (EEG), MRI, PET, and single-photon emission CT^4^. Invasive intracranial EEG (iEEG) recording using either depth electrodes (stereoelectroencephalography (SEEG)) or grid and strip electrodes (electrocorticography (ECoG)) is required in cases where non-invasive approaches are unable to localize the EZ with reasonable certainty^5^. Invasive measures, like iEEG, offer additional information for surgical planning, including improved temporal resolution of neural activity and non-seizure (interictal) biomarkers of the EZ^6,7^. However, even with these additional modalities, rates of seizure freedom following surgery remain relatively low with only 34% to 74% of patients (median of 62.4%) achieving long-term seizure freedom, according to a large meta-analysis^8^. This statistic may in part be accounted for by the difficulty of qualitatively demarcating the EZ in a complex ictal network, especially if there is no structural lesion on MRI^7^. Intracranial EEG monitoring exposes patients to risk from surgery, precipitated seizures—which may be more severe than typical epileptic events—and a weeks-long stay in the epilepsy monitoring unit, waiting for seizures. The need to develop more effective techniques for localizing the EZ is evident^9,10^.

The theory that epilepsy arises from a disordered brain network guides quantitative approaches to planning for invasive therapies^11^. Generically, these approaches consist of modeling normative brain activity by constructing a reference network—where regions of interest (ROIs) are the “nodes” and correlations of signals between ROIs are the “edges,” These models highlight abnormalities in a given patient’s brain network, and have shown promise for identifying the EZ^12–14^. An implementation of this method by Bernabei et al., which used a multicenter cohort of normative iEEG to map surgical targets, achieved an area under the receiver operator characteristic curve (AUC) of 0.77 in localizing seizure generators, using a combination of single-channel and inter-regional features^12^. A major limitation of the study however was that only awake iEEG recordings were used to construct the reference atlas, omitting potentially useful data obtained during sleep.

There is a well-established bidirectional relationship between epilepsy and sleep. Sleep disturbances are associated with the occurrence of nighttime seizures and also with the presence of epilepsy itself^15,16^. Recordings of electrical activity during sleep can reveal biomarkers (such as interictal epileptiform discharges) sometimes not apparent during wakefulness, which are important in surgical planning^17^. Seizures and epileptic discharges preferentially occur during non-REM sleep (NREM) as opposed to wakefulness or REM, a phenomenon that may be explained by the synchronizing effect of high-amplitude slow waves during NREM^18–22^. Support vector machine (SVM) models trained by Klimes et al. using univariate and bivariate features from iEEG recordings of stage N2 and N3 sleep localized the EZ better than those trained using REM and awake recordings^23^. Other work in scalp EEG has found that interictal epileptiform discharges (IEDs) are more concordant with the seizure onset zone in REM sleep compared to NREM sleep^24^. We hypothesize that an atlas of normative interictal iEEG during sleep would improve the accuracy of localizing the EZ by potentially identifying seizure-driving regions that are dormant during wakefulness, revealing NREM or REM features with high prognostic value, and better characterizing the overall evolution of the epileptic network and its circadian rhythms. Additionally, a tool for appraising interictal iEEG data during sleep would reduce the burden on patients and institutions by shortening EMU stays and reducing the number of epileptic events per patient before surgery.

In this study, we developed a normative iEEG atlas that includes data from both awake and sleep stages. Building on our previously validated methods^12^ to construct normative iEEG atlases for wakefulness, we included N2, N3, and REM sleep stages to define normal iEEG activity. We hypothesize that (i) abnormality features captured during NREM sleep would better localize the EZ than those captured during REM and wakefulness and (ii) awake and sleep abnormalities are associated with comorbid neuropsychiatric disorders. We have previously proposed that novel methods for optimizing epilepsy surgery based upon large, multi-centre datasets may streamline and improve presurgical localization of epileptic networks and tertiary care^12,25^. Towards this aim, we publicly release all iEEG data and code to foster ongoing collaboration in this space. Overall, we aim to use iEEG data captured from different stages of vigilance to improve localization of the EZ and likelihood of postoperative seizure freedom in patients with DRE.

## Materials and methods

### Subject information

We retrospectively analyzed a cohort of 108 subjects (*n*_female_ = 58) with DRE. Each patient had undergone implantation with subdural grid-and-strip electrodes, stereotactic depth electrodes, or a combination of both. Twenty-nine subjects had primarily ECoG grids and 76 subjects had primarily SEEG depth electrodes. From this data, we applied the following inclusion criteria: 1) subject completed resection or ablation surgery, 2) subject was evaluated at least 2 years after surgery by a board-certified epileptologist, 3) subject received a post-operative T1 MRI from which the resection zone could be manually segmented, 4) subject did not have a resection or ablation prior to iEEG implantation, and 5) subjects had at least 2 different sleep stage classifications over a 12 hour night-time sleep staging period. Of the 108 subjects, we progressively applied all criteria and 30 subjects were ultimately included in the study. 65 of the original 108 subjects were assessed for seizure outcome at least 2 years and did not have prior neurosurgery. 56 of those 65 had quality checked electrode localizations. Finally, 30 of those 56 had usable post-operative imaging to draw resection masks and map gray matter electrode contacts to the resection zone. The demographics and other clinical factors for the patient cohort are summarized in Table 1.

### iEEG data collection

Intracranial electrode configurations (Ad Tech Medical Instruments, Racine, WI) consisted of linear depth electrodes (1.1 mm in diameter with 5 mm spacing between contacts), and linear cortical strips and two-dimensional cortical grid arrays (2.3 mm in diameter with 10 mm spacing between contacts). Recording sampling rates varied from 256-1024 Hz and signals were referenced to an electrode distant from the suspected seizure sites, typically located in the medullary cavity of the skull.

For sleep classification, we selected one continuous 12-hr night-time period per patient (8:00 PM - 8:00 AM local time) that occurred at least 24 hours after the onset of recording (to account for implantation and anesthesia effects) and did not contain any clinical seizures. Preprocessing included removing scalp and EKG channels and bipolar re-referencing of all intracranial channels. We applied a previously validated sleep staging algorithm, SleepSEEG, to a total of 108 subjects with available metadata and 12-hour iEEG recordings that met this criteria^26^. This yielded classifications of each contiguous 30-second interval within each 12 hour recording into wake, stage N1 sleep, stage N2 sleep, stage N3 sleep, or REM sleep. N1 sleep classifications were omitted from all further analyses due to poor classification confidence from the SleepSEEG algorithm. Segments where one stage was exclusively classified for over an hour or that contained artifacts were excluded due to noise. From the remaining classifications, we identified the 30-second interval for each sleep stage that was at the midpoint of the longest continuous classification of that stage. This method ensured that any uncertainty in sleep stage classification was minimized when selecting representative clips.

Awake segments were selected outside of the 12-hr nighttime period and met the following criteria: 1) artifact free, 2) 2 hours before seizure onset, 3) 2, 6, and 12 hours before subclinical, focal, and generalized seizures, respectively, 4) minimal interictal spikes, and 5) at least 24 hours after implantation (**Figure 1A**).

To all awake and sleep recordings, we first eliminated artifact dominated channels with a custom artifact detector. Briefly, each 1-s window for a given recording was checked for disconnection and overwhelming high frequency and amplitude noise. If more than 3-s of a channel were marked as artifact, the entire channel’s recording was omitted. Channels were montaged using bipolar re-referencing such that the differences between adjacent contacts were taken (e.g. electrode contacts LA01, LA02, LA03, LA04 to LA01-LA02, LA02-LA03, LA03-LA04). Data were then filtered for powerline noise using a second-order infinite impulse response filter with quality factor 100 at 60 Hz and harmonic frequencies 120 Hz and 180 Hz. Zero phase filtering was applied to prevent phase delay. All data were then downsampled from their original sampling frequency to the lowest factor above 200 Hz (e.g. 1024 Hz was downsampled to 256 Hz and 500 Hz was downsampled to 250 Hz). This protocol ensured consistency and minimized computational cost since downstream features only used signals between 1 Hz and 80 Hz. The mean of each channel in each recording was subtracted from the recording to center each channel around 0 and remove DC offset.

To increase the sampling of the normative atlases, data from the MNI Open iEEG Atlas were combined with data from the HUP iEEG cohort. This data consists of 1772 channels with normal brain activity from 106 subjects from the Montreal Neurological Institute and Hospital, the Grenoble-Alpes University Hospital, and the Centre hospitalier de l’Université de Montréal^13,27,28^. We downloaded this data from the publicly available repository and clipped recordings to 30-s. These data include channels from normal brain regions in a cohort of patients undergoing iEEG monitoring during evaluation for epilepsy surgery.

### Electrode and surgical target localization to custom brain atlas

Each electrode contact in the HUP cohort was localized using the iEEG-recon tool^29^. Electrodes were annotated on post-iEEG-implantation CT scans, post-implant scans were linearly registered to a pre-implant T1-weighted MRI scan, and the transform matrix was applied to electrode coordinates. We ran Freesurfer *recon-all* on the T1-weighted MRI to register the Desikan-Killiany-Tourville (DKT) atlas to each subject’s native pre-implant MRI space^30^. As the channels were re-referenced using a bipolar montage, we computed the midpoint between the two original contacts as the localization of the bipolar “contact”. We constructed a 2-mm radius sphere around each contact and computed the overlap of the sphere with DKT atlas regions. Each contact was then mapped to the region of interest (ROI) with the highest percent overlap. Each bipolar channel in the MNI cohort contained metadata on the ROI from which it was recorded from using a custom 38-ROI atlas (see Supplementary Materials).

To identify the resection zone, we first collected post-procedure MRIs in subjects who received resection or ablation procedures (*n* = 44). When possible, we selected a 1mm isotropic T1-weighted MRI scan, and in subjects where this data was missing, we opted for axial T1-weighted MRI, sagittal T1-weighted MRI, and axial post-contrast T2-weighted MRI. We used the Convert3d command-line tool to reslice anisotropic scans to isotropic resolution. We used the semi-automatic segmentation tool in ITK-SNAP to label resection cavities and ablated tissue in the isotropic resolution post-procedure MRIs^31^. Finally, we linearly registered the post-procedure MRI to the pre-implant MRI space and applied this transformation to the annotated resection or ablation. Contacts that were inside the resection or ablation zone or less than 5mm from the margin by Euclidean distance were labeled as being within the resection zone.

To ensure sufficient electrode sampling of each ROI, we defined a custom atlas in which we condensed anatomically and functionally similar ROIs from the DKT atlas and mapped MNI atlas ROIs to our custom atlas. This resulted in 40 ROIs (20 in the left hemisphere, 20 in right hemisphere) to which each bipolar channel could be mapped (**Figure 1B**). A mapping table is provided in Table S1.

### Univariate and bivariate feature computation

To represent neurophysiological epileptogenic network properties across different frequencies of neural oscillations, we computed a set of univariate and bivariate features from each iEEG channel across both HUP and MNI datasets for each sleep and awake recording. We computed spectral power in 5 canonical frequency bands and broadband: delta — 1 to 4 Hz; theta — 4 to 8 Hz; alpha — 8 to 12 Hz; beta — 12 to 30 Hz; gamma — 30 to 80 Hz; and broad — 1 to 80 Hz. First, we estimated the power spectral density using Welch’s method^32^ over 2-s windows with 1-s stride using a Hamming window and computed the mean of the periodogram. To obtain non-negative areas under the curve, we log-transformed the power spectrum and offset all log-transformed powers to be non-negative. For each frequency band, we integrated powers within that band using the composite Simpson’s rule. We transformed the canonical frequency bands (all except broadband) to relative bandpower by dividing power within that band by power within all bands. Thus, all canonical frequency band powers summed to unity (**Figure 1C**).

To complement the univariate features, we computed magnitude squared coherence for each pair of channels within each recording. We computed coherence by applying a Hamming window over 2-s windows with 1-s stride. To condense coherence measurements to a scalar for each frequency band, we computed the mean coherence for canonical frequency ranges and across all bands for the broadband coherence feature. Univariate and bivariate features were independently computed for each sleep and awake stage recording (**Figure 1C**).

### Construction of normative atlases

We combined the computed features for each bipolar channel with its ROI assignment in the custom atlas. Consistent with previous work, we defined an abnormal channel as being either within the seizure onset zone (SOZ), the irritative zone (greater than one spike per hour), or the resection zone^12,13^. Channels that did not meet any of the criteria for abnormality were deemed normal and combined within each ROI of the custom atlas. Under this definition, the HUP dataset included 513 clinically normal channels and 7219 abnormal channels from 30 subjects. All MNI Open atlas channels (*n* = 1772) were included in the normal channel dataset. The same normal and abnormal classification criteria were applied for all sleep and wake stages.

Univariate and bivariate features for each bipolar contact were transformed into *z-*scores relative to normal channels within the same ROI. For one univariate feature from a given channel, we determined the normative distribution by assembling that feature from all other channels that were deemed normal and not sampled from the same subject to preserve a leave-one-subject-out (LOSO) methodology. We then used the mean and standard deviation of the normative distribution to transform the univariate feature from a relative bandpower measure to a *z*-score, thus normalizing the feature to the ROI. This procedure was repeated independently for all 6 univariate features in each channel and each wake/sleep stage in our dataset.

Bivariate features were similarly referenced to the normative atlas. Each bivariate feature, measured between ROI A and ROI B, were *z*-scored against all normative coherence features between ROI A and ROI B. Normative bivariate features were defined as those where a subject had sampling of ROIs A and B, and the contacts in those ROIs between which coherence was calculated both met the above criteria for being deemed normative. We again excluded features from the test subject to preserve leave-one-subject-out methodology. The mean and standard deviation of these normal features measured between ROIs A and B were used to transform the bivariate features into *z*-scores. To project bivariate *z*-scores from each pair of channels to each channel, we replicated methods from previous literature^12^. We estimated each bipolar contact’s abnormality by computing the 75th percentile of all bivariate *z*-scores projecting from that contact. Thus, each bipolar contact was represented by 6 univariate *z*-score features and 6 bivariate *z*-score features (**Figure 1D**).

### Statistical analysis

Across all analyses, we opted for non-parametric statistical tests over parametric counterparts to avoid making assumptions about the underlying distribution of features across channels, ROIs, and sleep stages. Moreover, some regions had small sample sizes, for which non-parametric statistics are preferred^33^. The Wilcoxson test was used for paired sample tests and the Mann-Whitney test was used for non-paired tests. We applied Holm-Bonferroni correction to correct for multiple comparisons when applicable. When interpreting effect sizes, we adopted the Cohen’s *d* convention where 0.2, 0.5, and 0.8 represent small, medium, and large effects, respectively^34^.

To predict abnormality, we developed a classifier pipeline to predict channels that were deemed normal and those that were most abnormal: contacts that were within the resection zone in subjects who had Engel 1A or 1B outcome at 24 months following surgery. We first split normal and abnormal channels across both the HUP (both normal and abnormal) and MNI (normal only) datasets using stratified group 5-fold cross-validation where groups were determined based on the subject from which the channel was recorded and each fold was stratified to preserve the ratio of abnormal and normal channels. We trained a model pipeline on the 12 univariate and bivariate *z*-score features. The model pipeline consisted of three sequential modules: 1) Synthetic Minority Oversampling Technique (SMOTE) applied to the minority (abnormal) class, 2) random undersampling applied to the majority (normal) class, and 3) a random forest classification model with 100 estimators. A pipeline was independently fit to normal and abnormal channels in each group of four folds and applied to all channels in the fifth fold. For subjects that were not in any of the training folds, those that were not candidates for surgery or had poor surgical outcome, we retrained the pipeline on all folds of data and predicted abnormality on this subset of subjects. We ensured that predictions from a model had never seen data from that subject and maintained consistent cross-validation folds across all subsequent analyses (**Figure 1E**).

Channel-level abnormality predictions were compared to clinical hypotheses of the EZ. We first registered manual segmentations of the resection or ablation zone into each patient’s pre-operative space and mapped each electrode contact to belonging within or outside the surgical target. If a contact was within 5 mm of the resection or ablation boundary, it was marked as targeted, and all other contacts were marked as spared. We then computed the area under the precision-recall curve (AUPRC) where the true values were binary labels of targeted or spared, and the predicted values were predicted abnormalities for each electrode contact. AUPRC is preferred for imbalanced classes, as typically more contacts were spared than targeted. Thus, each model’s channel-level predicted abnormalities were summarized at the patient-level with an AUPRC metric, where higher (lower) AUPRCs indicated larger (smaller) differences in predicted abnormalities between targeted and spared contacts (**Figure 1E**).

**Figure 1.**
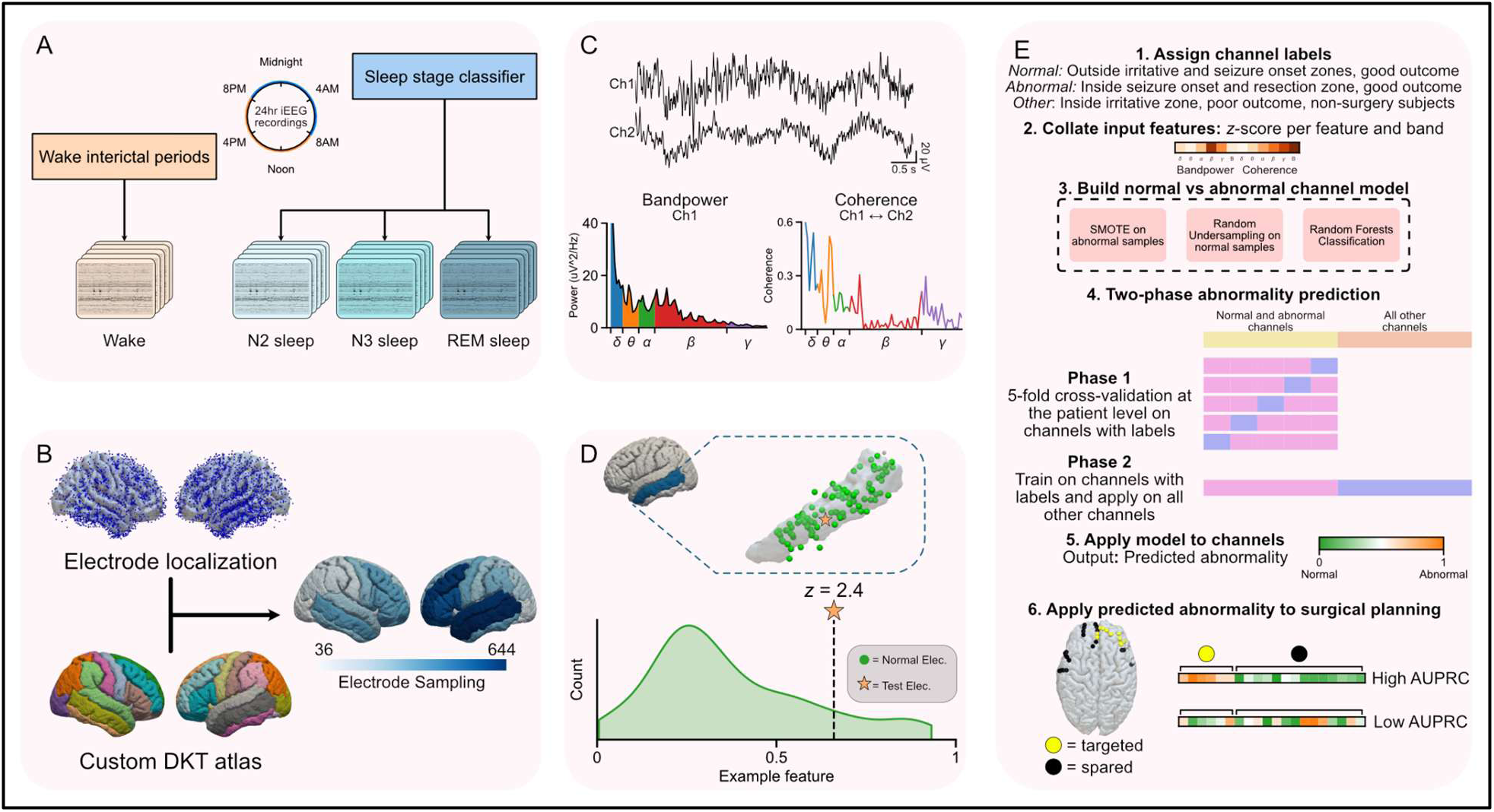
Methods overview. A) Intracranial EEG epochs are extracted from awake, interictal periods, and sleep periods. Sleep periods are obtained from applying a 12-hour night-time (8:00PM to 8:00AM) recording to a sleep stage classifier. Epochs that have the highest confidence classification as N2, N3, and REM sleep are extracted as representative iEEG from that sleep stage. B) Electrodes are localized using *iEEG-recon* and mapped to an aggregated DKT atlas. C) From each epoch, we extracted bandpower (univariate) and coherence (bivariate) features. D) Each feature was transformed into *z*-scores based on the other feature values from that atlas region that were deemed normal. E) We predicted the abnormality of each electrode contact by training a random-forest model on the *z*-scores for univariate and bivariate features to predict normal and abnormal channels in good outcome patients. We applied this model to all patients, and aggregated patient-level metrics by computing the area under the precision-recall curve (AUPRC) of the resection and non-resection zones.

## Results

### Multi-center interictal iEEG from drug-resistant epilepsy patients

To best capture epileptic abnormalities, we selected a heterogeneous study cohort of DRE patients who presented to our tertiary-level epilepsy center. Thirty subjects met our inclusion criteria for this study. Surgical targets included temporal, mesial temporal, and frontal structures. Subjects were followed for up to 24 months after surgery, and seizure freedom was evaluated on the Engel scale by a board-certified epileptologist. Eighteen of the 30 subjects sustained seizure freedom across two years, and 12 subjects were not seizure free. Seizure outcome and detailed demographic information for each subject are provided (**Supplementary Table 1**).

Recognizing the limitations of a single-center approach and to best approximate normal brain electrophysiology, we supplemented our data with data available from the MNI Open iEEG Atlas^13,27,28^. This consisted of normal awake and sleep iEEG from four epilepsy centers. From our data and the MNI Open Atlas, we selected up to 5 minutes of data per sleep and awake stage, limited by detection of sleep with the sleep stage classifier. Following the extraction of bandpower and coherence features, we observed medium effect sizes in features between data collected from HUP and the MNI Open iEEG database, suggesting that some multi-center features could be harmonized (**Supplementary Figure 1**).

**Table 1.**

|  |  | Engel 1 @ 2yrs<br>(n = 18) | Engel 2+ @ 2yrs<br>(n = 12) | Chi-squared test |
| --- | --- | --- | --- | --- |
| **Target** | Frontal | 2 | 3 | n.s. |
|  | Other | 2 | 0 |  |
|  | Temporal | 14 | 9 |  |
| **Laterality** | L | 6 | 8 | n.s. |
|  | R | 12 | 4 |  |
| **Implant** | ECoG | 9 | 2 | n.s. |
|  | SEEG | 9 | 10 |  |
| **MRI Lesion Status** | Lesional | 7 | 2 | n.s. |
|  | Non-Lesional | 11 | 10 |  |
| **Age** | 10-19 | 1 | 0 | n.s. |
|  | 20-29 | 7 | 6 |  |
|  | 30-39 | 5 | 5 |  |
|  | 40-49 | 4 | 1 |  |
|  | 50-59 | 1 | 0 |  |
| Has N2 | FALSE | 1 | 0 | n.s. |
|  | TRUE | 17 | 12 |  |
| Has N3 | FALSE | 2 | 1 | n.s. |
|  | TRUE | 16 | 11 |  |
| Has R | FALSE | 8 | 4 | n.s. |
|  | TRUE | 10 | 8 |  |
| Has W | TRUE | 18 | 12 | N/A |
| Engel 1 @ 1yr |  | N/A | 3 | N/A |
| Engel 2+ @ 1yr and Engel 1 @ 2yr |  | N/A | 1 | N/A |

### SleepSEEG generalizes to external datasets

Intracranial EEG recordings during sleep measure unique neurophysiological dynamics compared to wakefulness. These include sleep spindles^35^, sigma band spectral power, and K-complexes^36^. To isolate recordings during various sleep stages, we performed sleep staging using the publicly available SleepSEEG tool^26^. SleepSEEG successfully ran on 107 subjects. Fourteen of the subjects were excluded from consequent analyses for having poor classification with SleepSEEG, and > 90% of the 12 hour period was classified as being in one stage for these subjects. For all other subjects, we observed patterns of multiple sleep cycles and most awake classifications occurring at the beginning and end of the 12-hour sleep staging period (**Figure 1A**). We validated SleepSEEG findings by computing the alpha-delta spectral ratio on each channel. Across all sleep stages, we observed global differences in sleep and awake alpha-delta ratio (**Figure 1B**). In accordance with our prior hypotheses, we observed higher alpha-delta ratio in awake recordings when compared to sleep stages (W vs. N2: *W* = 42, *p*_Holm-Bonferroni_ < 1e-4; W vs. N3: *W* = 3, *p*_Holm-Bonferroni_ < 1e-4, W vs. R: *W* = 0, *p*_Holm-Bonferroni_ < 1e-4) (**Figure 1C**). We additionally observed higher alpha-delta ratio in N2 sleep and REM sleep than N3 sleep (N2 vs. N3: *W* = 0, *p*_Holm-Bonferroni_ < 1e-4; R vs. N3: *W* = 26, *p*_Holm-Bonferroni_ < 1e-4) (**Figure 1C**). Given the evidence that interictal epileptiform spikes occur at higher rates during sleep compared to wakefulness^19,37,38^, we used a validated spike detector to estimate spike rates during each of the stage classifications. We observed monotonically increasing spike rates across REM sleep, wakefulness, N2, and N3, in ascending order, concordant with previous findings (**Figure 1D**). Thus, we validated an external tool on our dataset and employed it for automatic sleep staging from SEEG and ECoG, as described above.

**Figure 2.**
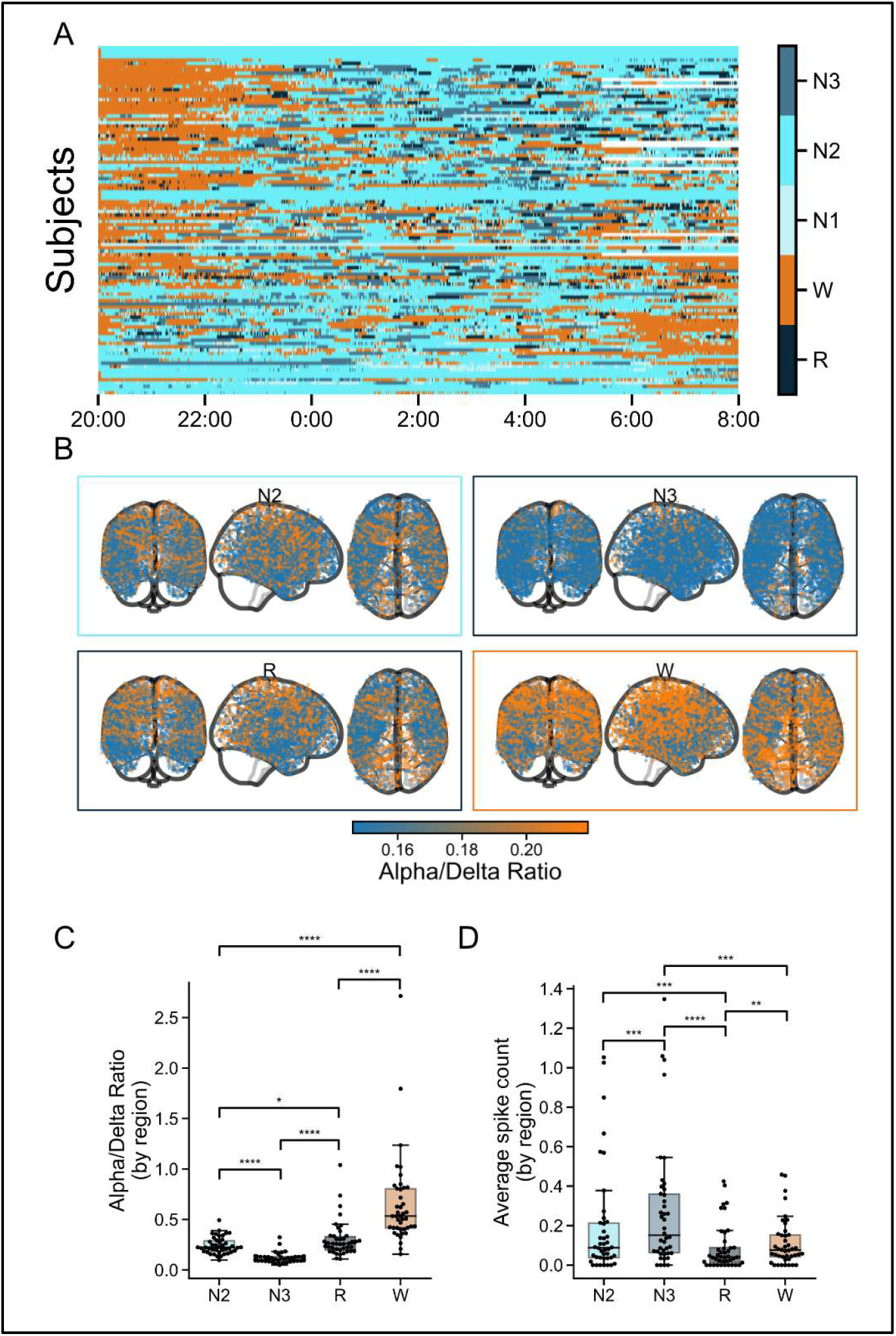
Analyzing and validating SleepSEEG. A) We applied SleepSEEG to 108 subjects who had at least 12 hours of nighttime EEG without clinical seizures at least 24 hours after implantation. 30 second segments were classified as awake, REM, N1, N2, or N3 sleep with probabilities from 0 to 1. B) Alpha-delta ratio computed on each extracted epoch in each sleep stage revealed global differences between sleep and wake stages. C) Alpha-delta ratio was highest in awake and lowest in N3 (Wilcoxson signed-rank test; *p_Holm-Bonferroni_* < 0.05 for marked pairs). D) We tested differences in average spike count in the extracted awake and sleep epochs and observed higher spikes in N3 than N2, REM, and wake (Wilcoxson signed-rank test; *p_Holm-Bonferroni_* < 0.05 for marked pairs).

### Epileptic abnormalities are detected across sleep and wakefulness

To detect abnormal channels, we built a model that used channel *z-*scores to predict whether that channel was surgically targeted or not in subjects with good outcome (Engel 1A or 1B at 24 months). This model was cross-validated with LOSO, and we generated abnormality scores from 0 to 1 for each channel based on the model’s predicted epileptogenicity with 1 being abnormal. We estimated abnormality in all subjects, including those with poor outcomes. This model condensed each channel’s *z-*scores from every band and both bandpower and coherence features into a single abnormality score, where values closer to unity represented channels with higher predicted abnormality and values closer to zero represented channels with lower predicted abnormality. We evaluated cross-validation performance by aggregating validation set predictions before computing the area under the receiver operator curve (AUC). Model performance was significantly better with data from REM sleep (AUC = 0.86, 90% CI: [0.81, 0.90]) than N2 sleep (AUC = 0.72, 90% CI: [0.67, 0.77]), N3 sleep (AUC = 0.70, 90% CI: [0.64, 0.75]), and wake (AUC = 0.69, 90% CI: [0.63, 0.74]) (**Figure 2A**) (Delong’s test; *p*_Bonferroni_ < 0.01 for REM & N2, REM & N3, and REM & awake).

As iEEG dynamically measures brain activity, we expected that short recordings in each sleep stage may vary in their capability to predict epileptogenicity. We selected up to 5 minutes of data per wakefulness stage and independently computed features, *z*-scores, and predicted epileptogenicity for each 30-s non-overlapping segment. We measured the difference in predicted abnormality for each channel for each shortened recording compared to the full recording and found comparable levels of error for each sleep stage that reduced as we approached 5 minutes of recording (**Figure 3B**). Importantly, the variance of predicted abnormality decreases as the amount of data used in the model increases, suggesting that the features converge as more data is included. Though the difference in the predicted abnormality was small (on the order of 0.01 out of 1), the decrease in variance as more data is included suggests that at least 5 minutes of iEEG data should be used for abnormality analysis.

To increase the interpretability of the models, we computed feature importance on each of the four models. We fit the base model on data from the entire training set, performed permutation testing on each feature independently, and obtained the decrease in accuracy as a marker of feature importance. We elected to perform permutation testing since many of the features exhibited multicollinearity (**Supplementary Figure 2**). Among the 12 features (6 canonical bands across bandpower and spectral features), we found that delta and theta bandpower are among the least important features and broadband coherence and gamma coherence are among the most important features (**Figure 2C**).

We then repeated our analysis across a series of thresholds for what we deemed a good outcome after surgery. This threshold filtered which contacts we considered normal—normal contacts were selected from good outcome subjects only—and affected the training labels for the random forest models that predicted epileptogenicity. We evaluated thresholds of good outcome as Engel 1A, 1A + 1B and 1A-D at 12 months and sustained through 24 months. The AUC of predicting normal and abnormal channels was best in REM sleep and awake at 12 month outcomes, and best in awake at 24 month outcomes (**Supplementary Figure 3**). With most stages and good outcome criteria, we observed good test-set performance of normal versus abnormal classification (median AUC = 0.70, IQR = 0.11). For downstream analyses, we fixed the criteria for good outcome as subjects with at least 24 months of follow up after surgery and Engel 1 outcome at 12 and 24 months.

**Figure 3.**
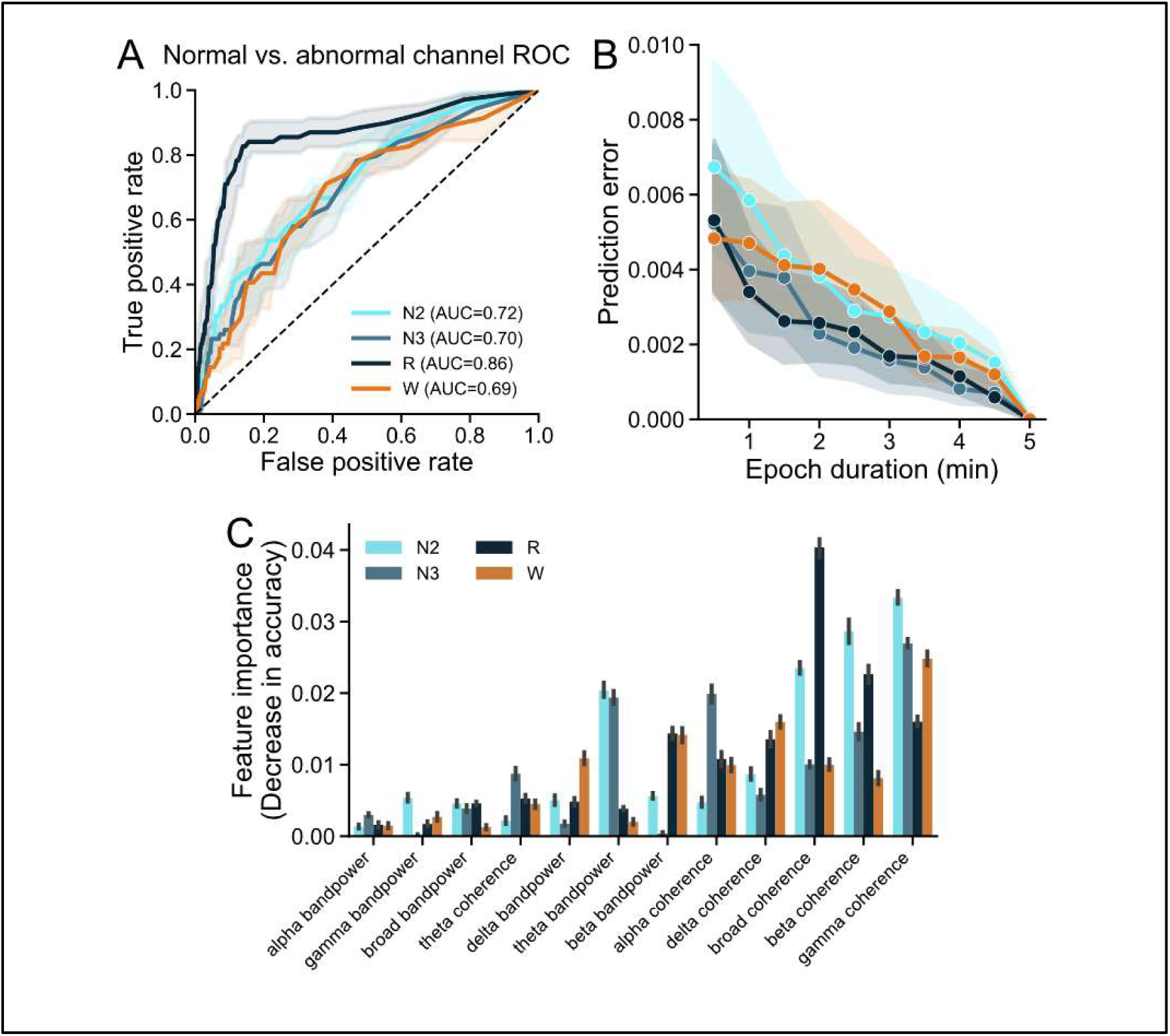
Computing predicted abnormality across sleep and wakefulness. A) We applied a cross-validated model on *z*-score features to predict normal and abnormal channels in good outcome patients using gold-standard resection labels. All models achieved good performance at predicting normal and abnormal channels and we obtained abnormality scores for each channel in good and poor outcome subjects that ranged from 0 to 1. Confidence intervals are obtained using the 5th and 95th percentile AUC values when bootstrapping the normal and abnormal labels over 100 iterations. B) We determined how stable the prediction of each sample was by computing the absolute difference in predicted abnormality from 5 minutes of recording to predicted abnormality from a shorter epoch duration. C) We computed feature importance for each model using permutation-based feature importance. Decrease in accuracy is reported for each iteration of omitting a given feature.

### Sleep abnormalities localize resection and laser ablation targets

The clinical hypothesis of epilepsy surgery necessitates a focal EZ within the network, and that targeting this EZ will improve seizure control. We tested if the abnormalities detected in sleep and wakefulness localize the EZ, and hypothesized that patients with seizure control at 24 months after surgery would have higher and concentrated abnormalities in the surgically targeted tissue compared to surgically spared tissue. Conversely, those with poor seizure control were hypothesized to have poor distinction between abnormalities within and outside the resection zone. To test this hypothesis, we computed the area under the precision recall curve (AUPRC) for each set of sleep/wake stage abnormality scores from each patient. The true labels were set to channels recording from surgically targeted or spared tissue, and the predicted scores were set to the abnormality scores. AUPRC is sensitive to low sample sizes and class imbalance, which was ideal for our case where a majority of channels were surgically spared. A higher AUPRC metric indicated better separation in abnormality scores between surgically targeted and spared tissues, and a lower AUPRC metric indicated more homogenous abnormalities within and outside the surgically targeted regions. We separated subjects into two cohorts—those with good (Engel 1) and poor (Engel 2+) outcomes at 24 months—and computed the effect size between the two cohorts. The area under the precision recall curve between resected and non-resected channels was significantly higher in good outcome patients than those with poor outcome for models trained on data from N2 sleep after correcting for multiple comparisons (**Figure 4**). Models from N2, N3, and awake stages had large effect sizes.

**Figure 4.**
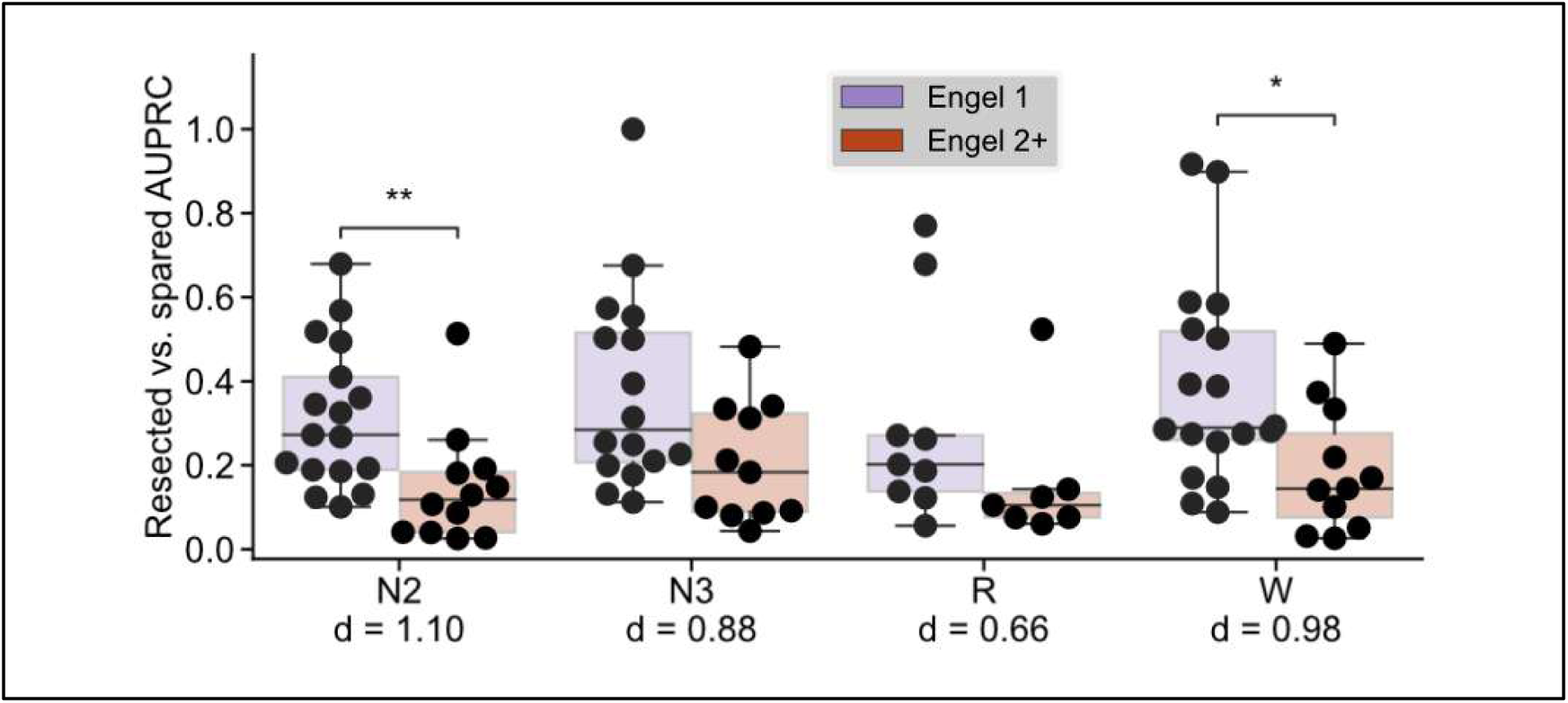
Computing patient abnormalities. For each subject we computed the area under the precision-recall curve (AUPRC) using the predicted abnormalities as the features and the binary labels of surgically targeted and spared. Subjects with Engel 1 outcome at 2 years had higher AUPRCs than those with Engel 2+ outcome in N2 sleep and wake (Mann-Whitney U test; *U_N2_* = 169, *p_N2_* = 0.003, *U_W_* = 154, *p_W_* = 0.015). Cohen’s *d* effect sizes are also reported.

### Combining sleep and awake features improves epileptic network localization

As awake and sleep data provide complementary information on brain activity, we replicated our prior analyses while testing combinations of wakefulness and sleep-based feature combinations for predicting normal and abnormal channels. We found that combining awake and N2 or N3 features improved classification of normal and abnormal channels as determined by the area under the receiver operating curve beyond either stage by itself (**Figure 5A**). To understand how combining stages affected localization of the EZ, we computed the effect sizes between good and poor outcome subsets for each stage by themselves and combined. We observed improvements in the separability of good and poor outcome subject groups when combining N2 with wake and N3 with wake (**Figure 5B**).

To further investigate how predicted abnormalities compared with the surgical target, a proxy for the clinical hypothesis of the epileptogenic network, we highlighted the four examples from the model combining N2 sleep and wakefulness, which had the highest effect size (**Figure 5C**): a subject with good outcome and high AUPRC (**Figure 5D**), a subject with good outcome and low AUPRC (**Figure 5E**), a subject with poor outcome and high AUPRC (**Figure 4F**), and a subject with poor outcome and low AUPRC (**Figure 4G**). Subjects D and G agreed with our hypothesis that patients with good outcomes would have higher AUPRC than those with poor outcomes and subjects E and F disagreed with our hypothesis. Predictions for subject D correctly marked abnormalities in a frontal lobe epilepsy case where temporal lobe contacts were not predicted as abnormal. Subject E was bilaterally implanted and abnormalities were found in both temporal lobes whereas a procedure in the right temporal lobe was sufficient to achieve seizure control. This example suggests that there may be subregions of abnormal epileptic networks that are more critical to seizure generation than others, and that targeted interventions in these critical areas may be sufficient. It might also suggest regions of the epileptic network in different stages of epileptogenesis, with implications for seizure relapse in the future. Subject F was implanted with an ECoG grid across the left temporal lobe, of which most contacts were correctly predicted abnormal, though this patient was not seizure free. This finding raises the issue of sampling error, and the limitations inherent in iEEG and its restricted approach to mapping the brain only around implanted electrodes. Subject G had widespread bilateral abnormalities and received a targeted left mesial temporal lobe ablation. In cases such as this, it might be most useful to look at degrees of abnormality, or specific biomarkers in sampled regions to identify those areas most likely to participate in seizure generation.

**Figure 5.**
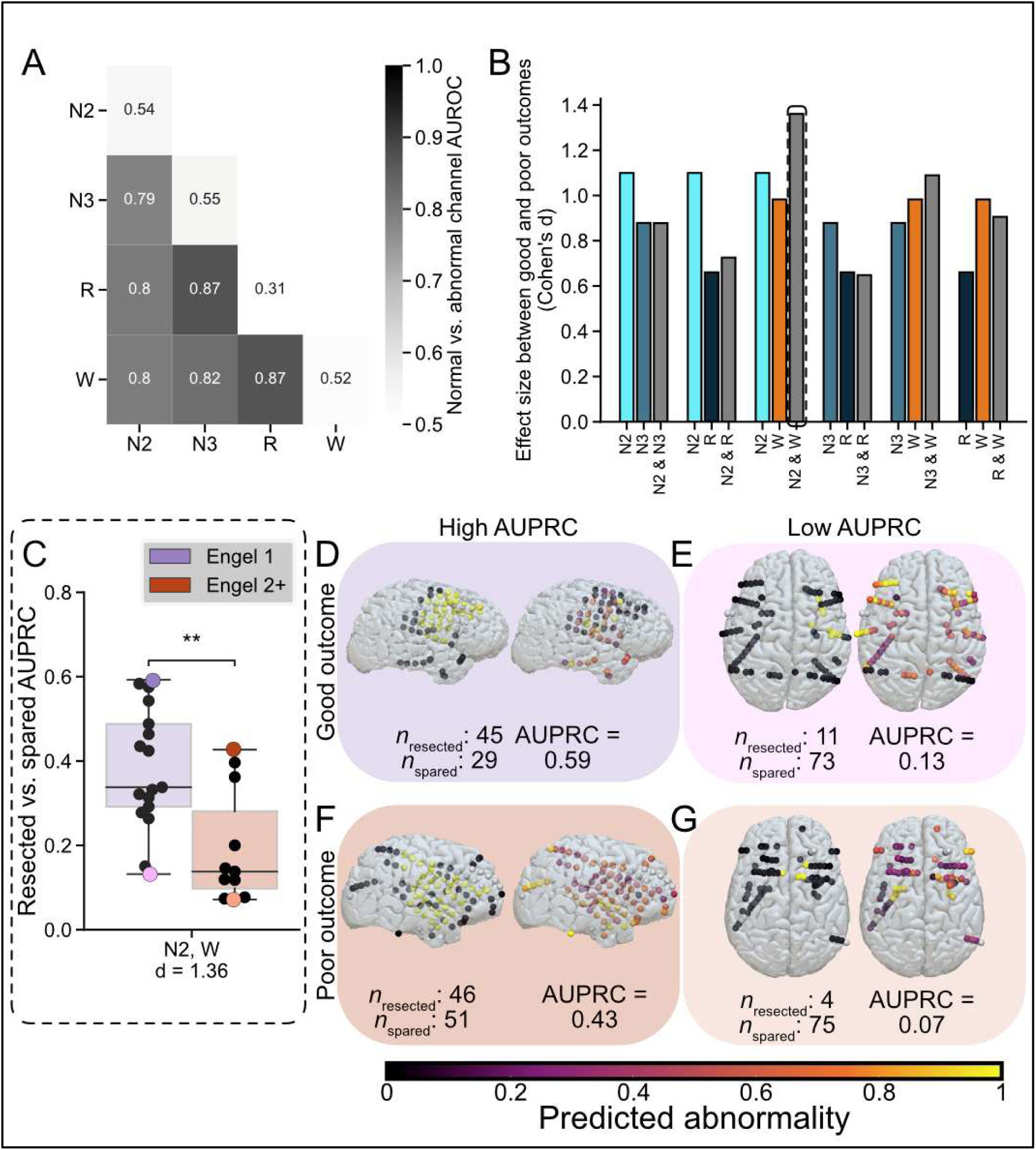
Combining sleep and wake abnormalities. A) Random forest models predicted normal and abnormal channels using combinations of sleep and wake stages (test AUC range = [0.79, 0.87]. B) We observed the biggest increase from univariate effect size between Engel 1 and Engel 2+ AUPRCs in combining N2 and wakefulness (AUPRC_N2_ = 1.1, AUPRC_W_ = 0.98, AUPRC_N2 + W_ = 1.36). C) The combination of N2 and awake features best separated Engel 1 and Engel 2+ cohorts, and we extracted four example subjects. D-G) We visualize resection masks and predicted abnormalities for subjects with high AUPRC and good outcome (D), low AUPRC and good outcome (E), high AUPRC and poor outcome (F) and low AUPRC and poor outcome (G).

### Awake and sleep abnormalities are associated with epilepsy comorbidities

Sleep quality has been linked to neuropsychiatric disorders, so we hypothesized that greater sleep abnormalities were associated with poor performance on neuropsychological testing that is conducted as part of routine epilepsy surgery planning. As part of clinical workup prior to intracranial monitoring, subjects completed at least one out of five different neuropsychological examinations: Full-Scale Intelligence Quotient (FSIQ), Verbal Comprehension Index (VCI), Perceptual Reasoning Index (PRI), Beck Depression Inventory (BDI), and Generalized Anxiety Disorder 7 (GAD-7). To represent the various neuropsychological tests while being parsimonious to avoid several multiple comparisons, we computed pairwise correlations in scores between the five tests. We found two clusters of tests, where cluster 1 consisted of the FSIQ, VCI, and PRI and cluster 2 consisted of the BDI and GAD-7 (**Supplementary Figure 4**). We selected the test from each cluster with the highest number of subject participants: the BDI and VCI.

Our hypothesis was that global abnormalities in each subject were associated with poor neuropsychological exam performance, and we summarized each patient’s degree of global abnormality by aggregating mean measures across all contacts. For each test, we fit a linear regression model where the predictors were neuropsychological performance test score and the regressors were mean abnormalities and interictal epileptiform spike rate in each stage. We selected interictal epileptiform spikes as a pre-machine learning model baseline. We hypothesized a positive correlation between BDI scores and iEEG features and negative correlation between VCI scores and iEEG features, where in both cases more severe neuropsychological scores are associated with more elevated iEEG features. Further, we hypothesized higher feature importance in the abnormality features than in spike rate.

Predicted BDI scores from the iEEG features approximated true scores moderately (**Figure 6A**; R2 = 0.63) and abnormalities across sleep and wakefulness had higher feature importance than spike rates. The most important feature was N3 abnormality which was positively correlated with BDI scores (**Figure 6B**). We observed similar performance with the VCI scores when approximating with iEEG features (**Figure 6C**; R2 = 0.57). NREM and REM sleep abnormalities were more important than wake abnormalities and sleep or wake spike rates (**Figure 6D**).

**Figure 6.**
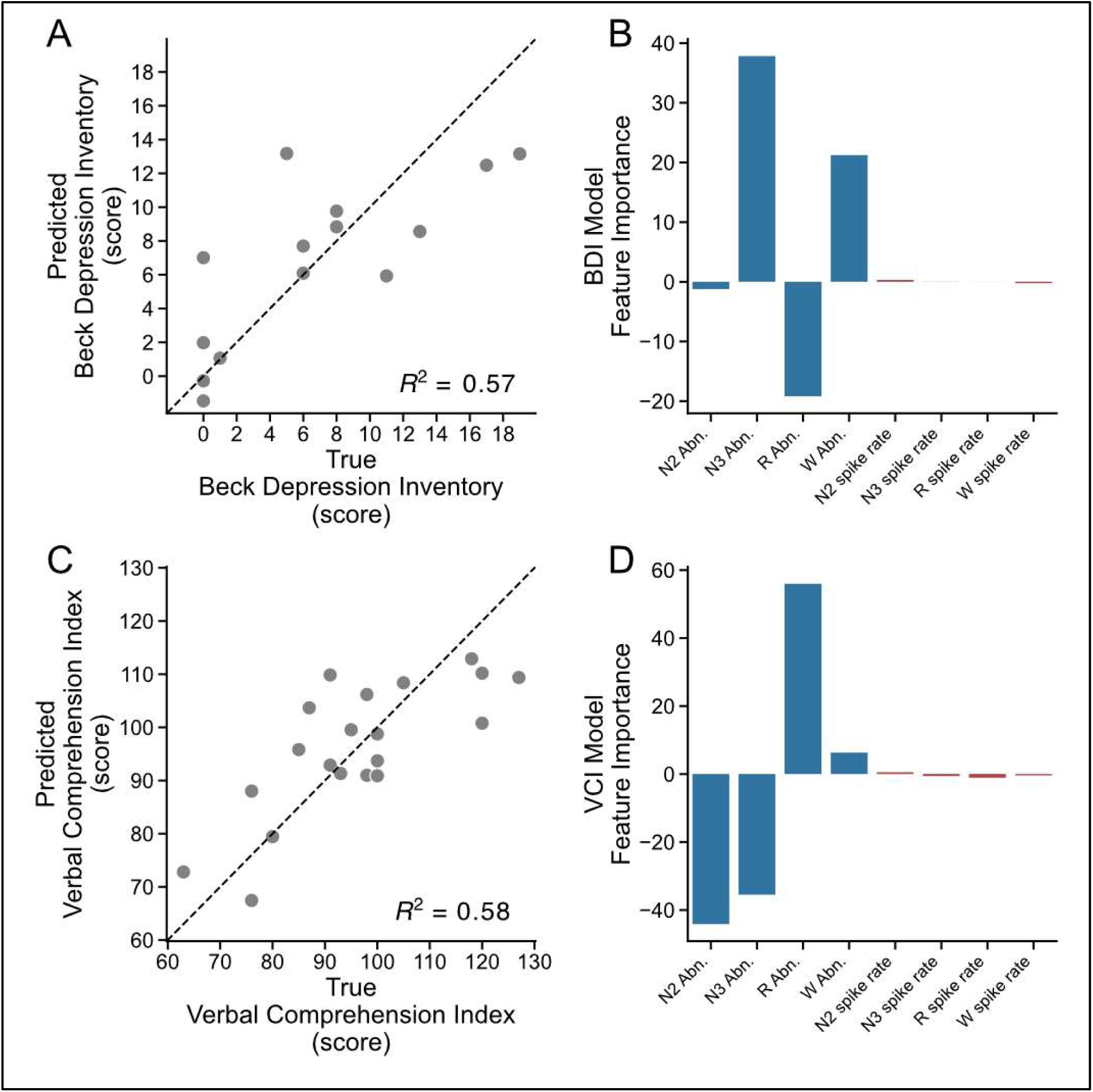
Associating sleep and awake abnormalities with neuropsychiatric exam performance. A) A linear regression model fit on Beck Depression Inventory (BDI) scores with N2, N3, REM, and wake median abnormalities and spike rates moderately approximated true scores (R^2^ = 0.57). B) Model coefficients revealed that abnormality features had higher magnitudes than spike features. C) Methods from A were replicated for the Verbal Comprehension Index (VCI) score (R^2^ = 0.58). D) Methods from B were replicated, and abnormality features had higher magnitude than spike features.

## Discussion

In this study, we sought to measure differences in models trained on data from different sleep and awake stages in identifying epileptogenic tissue from a cohort of DRE patients. The novel contributions of this study are 1) external validation of a sleep stage classifier for SEEG data, 2) construction of a normative atlas across sleep stages, 3) head to head comparison of EZ localization for parallel methods applied to wakefulness and sleep, and 4) comparison of iEEG biomarkers of abnormality with neuropsychiatric testing. We build upon previous findings and methods in the literature, and publish all code and data to continue advancing quantitative biomarkers for epilepsy^25^.

Localizing epileptiform tissues is the primary goal of pre-surgical workup in DRE^5,39^. We aimed to improve this procedure by computing abnormalities with respect to the anatomical region that an electrode contact is recording from. We used a machine learning model trained on both spectral and network features to determine an electrode contact’s likelihood of being epileptogenic and used this probability as a biomarker to localize epileptiform tissues. Importantly, there are two methods that we used to evaluate whether epileptiform tissue has been accurately identified by the machine learning models trained on sleep and awake stages. First, we computed channel-level predicted abnormalities and used validation set predictions to evaluate the models. Second, we aggregated channel-level predicted abnormalities for each subject and determined the degree of separation between channels that were targeted and spared during surgery. The first method treats each channel independently while performing channel-level classifications (quantified by AUC values) and the second method contextualizes channel-level predictions with the resection zone and uses univariate, non-parametric statistics to separate good- and poor-outcome subgroups^40^. While REM-based data best separated normal and abnormal channels, we found that NREM and NREM + Awake data aggregated across subjects best separated good and poor outcome patients.

Lack of consistent results between the channel-level and subject-level statistics may be a result of small and imbalanced datasets. Methods being developed to overcome the limited and sparse sampling of the seizure onset zone with iEEG should be integrated to increase confidence in model predictions^41,42^ at the channel level. At the patient-level, accounting for spatial sampling bias^43,44^ can improve the generalization of these methods. A larger, multi-centre dataset is also necessary to perform subject-level predictions of good and poor outcome based on abnormal biomarkers^45^.

### Detecting interictal abnormalities in sleep and combined awake and sleep recordings

We found the better separation of good and poor outcome groups on the models trained with data from N2 sleep compared to N3, REM, and awake stages. Further, combining N2 sleep and awake data results in better separation of good and poor outcomes in interictal recordings. Our findings are consistent with previous literature which has found that NREM sleep best localizes the EZ^23^. Our study also finds that a combination of awake and N2 sleep improves separation of good and poor outcome subgroups, suggesting that awake and NREM sleep provide complementary features that are sensitive to normal and abnormal electrophysiology. Applying permutation testing for feature importance revealed that high frequency network features (gamma and beta coherence) were most important across sleep and awake-based models. This suggests that features such as the synchronization of interictal spikes and sleep spindles across wakefulness and sleep are informative in identifying abnormalities. Moreover, high frequency network features have previously been found to be biomarkers of good and poor outcome subjects across surgery and neurostimulation^46^ and prior evidence has linked high frequency network features to short-range connections and found abnormally elevated high-frequency synchrony in local connections within epileptogenic cortex even when distal connections are normal^47^. Theories of gamma band entrainment, where repeated stimuli strengthen high frequency connections and modify the underlying functional connectivity^48,49^, can partially explain how gamma band activity and connectivity could be elevated in abnormal, resected tissues.

### Imprecise surgical treatment of epilepsy limits quantitative methods

Developing quantitative tools for iEEG to aid in epilepsy treatment is reliant on the “gold standard”: surgical procedures based on clinical hypotheses that are aggregated across EEG, medical imaging, pre-implant testing, and iEEG. With current standards of practice, only 30 to 40 percent of patients are seizure free after surgery long-term^1,50^. Our methods that use this gold standard for surgically targeted and spared brain regions are fundamentally limited by this low success rate. In this study, we tested different criteria for determining whether a subject had a good outcome and found that channel level predictions vary as the outcome threshold varies. This suggests that 1) even patients who have similar outcomes may have different disease courses and severities and 2) a bigger cohort is necessary to capture heterogeneities in epilepsy type that can overcome differences in outcome. We ultimately chose to deem subjects as having a good outcome as those who had Engel 1 scores at 12 and 24 months. Our choice of threshold was influenced by the possibility that other clinical decision making may influence seizure outcome after surgery, such as medication changes, and stable seizure freedom for at least 2 years is most likely due to the surgical procedure.

Therapies such as medications, neurostimulation, and neurosurgery are intended to reduce seizure burden. Measuring the effect of surgery on reducing seizure burden will require alternative measures of outcome such as seizure severity^51,52^ or improvements in quality of life. Recent studies that redefine the goal of invasive EEG monitoring can provide alternative gold standard labels for what can be considered a successful outcome from iEEG, such as determining the focality of the EZ and informing good and poor candidates for surgery^53^.

Analysis of patients in whom our atlas comparison did not predict outcome is instructive, particularly in light of the assumptions involved in surgical decision making. These patients demonstrate that identifying abnormal brain regions is not likely enough to specifically target surgery, as some patients respond to therapy in more limited regions. This suggests that more specific biomarkers of abnormality may be required to better target critical seizure generating nodes in the network for intervention. In addition, those patients with resected abnormalities who do not respond to treatment highlight the perils of limited brain sampling inherent in IEEG. Another point limiting all of these analyses is the absence of ground truth in assessing seizure-freedom, given low accuracy in reporting of seizures among patients due to postictal confusion/ amnesia, and our inability to detect subclinical seizures in the absence of continuous outpatient EEG monitoring^54^.

### Mapping iEEG abnormalities to epilepsy comorbidities

Though seizure control is the primary objective of epilepsy care, comorbidities also affect quality of life^55–58^. In this study, we sought to test our biomarkers’ sensitivity to neuropsychiatric exams that measure the severity of comorbidities. We found that interictal abnormalities and epileptiform discharges partially explain the variance in scores for patients who completed the Beck Depression Inventory and the Verbal Comprehension Index. Stratifying DRE patients by their risk of comorbidities and targeting brain regions that are affected by comorbidities could improve quality of life. Importantly, abnormalities in regions outside of the hypothesized seizure onset zone can affect comorbidities^59,60^. Other studies use subjects with intracranial EEG recordings to study cognitive functions, making the assumption that non-seizure onset zone recordings approximate normal brain activity and connectivity. Future work should contextualize findings on cognition with iEEG with measures of epileptogenicity, to ensure that findings generalize to healthy cohorts and uncover new mechanisms of cognition rather than highlight pathological tissue.

### Limitations and future work

Though our study shows the utility of an interictal iEEG biomarker for localizing the EZ, certain limitations must be noted. First, seizures have been shown to impact sleep stage duration and alter overall sleep architecture, which could possibly affect data collection^15^. We applied SleepSEEG by removing channels that were suspected to be epileptogenic to aim to minimize erroneous sleep staging due to epileptogenic tissues, yet it is possible that sleep staging errors occurred. To mitigate this limitation, we chose segments that were classified as N2, N3, and REM sleep with the highest probability, removed patients without apparent sleep cycles, and validated sleep stage classification with alpha-delta ratio and epileptiform spike detection. The ground truth labels for channels as normal and abnormal were limited to patients with good seizure outcomes after surgery. Notably, channels marked as abnormal were within the resection zone in good outcome patients. As a result, our model is tuned to detect abnormalities in DRE cases that benefitted from surgery, most of which were temporal lobe epilepsy cases. Future work should extend these analyses to patients who received alternative therapies such as neuromodulatory devices (e.g. responsive neurostimulator^46^ and deep brain stimulation) or continued medication which will require alternative definitions of what channels are normal and abnormal in these patients. We approximated normal electrophysiology in each anatomical region in wake and sleep, and the granularity of our parcellations were limited by the number of patients, number of electrode contacts, and spatial distribution of electrode contacts. Adding data from more subjects, especially from other level 4 epilepsy centers who may have different implantation practices, will ensure that we are more optimally approximating normative iEEG. We chose to analyze data from the first available non-seizure night of recording, and future work should explore dynamic changes in abnormality as anti-seizure medications are withdrawn^61^ and seizures are recorded.

While the relatively small number of patients analyzed in this study is a significant limitation, we share our data and code openly as part of an important international effort to build tools to improve patient care and accelerate research in epilepsy. Our primary goal is for others to continue to contribute data and improve these methods globally, which will strengthen these tools, and their impact^25^.

## Conclusion

Drug-resistant epilepsy patients undergo invasive EEG monitoring to localize abnormalities and plan for epilepsy surgery. From this data aggregated over subjects from multiple epilepsy centers, we constructed atlases of normal spectral and network features over wake and sleep. We found that quantitative predictions of abnormalities based on normative maps in wake and N2 sleep combined can localize the EZ and separate subgroups of patients with good and poor seizure control after 2 years. We also associated abnormalities with epilepsy comorbidities measured with neuropsychiatric testing. Future research should aim to test these methods in larger multicenter cohorts and prospectively, towards automated and precise localization of the EZ.

## Data Availability

Derivative data and analysis code are available on Pennsieve.io (https://discover.pennsieve.io/datasets/414) and Github (https://github.com/penn-cnt/Pattnaik-sleep-atlas).

https://github.com/penn-cnt/Pattnaik-sleep-atlas

## Acknowledgments

We thank Brian Prager for assistance with code organization and Darrell De Freitas for assistance with data organization.

