## Supplementary Materials for "Normative intracranial EEG highlights epileptic abnormalities across wakefulness and sleep"

### Materials and Methods

##### iEEG recording information

Intracranial electrode configurations (Ad Tech Medical Instruments, Racine, WI) consisted of linear depth electrodes (1.1 mm in diameter with 5 mm spacing between contacts), and linear cortical strips and two-dimensional cortical grid arrays (2.3 mm in diameter with 10 mm spacing between contacts). Recording sampling rates varied from 256-1024 Hz and signals were referenced to an electrode distant from the suspected seizure sites, typically located in the medullary cavity of the skull.

##### iEEG preprocessing

To all awake and sleep recordings, we first eliminated artifact dominated channels with a custom artifact detector. Briefly, each 1-s window for a given recording was checked for disconnection and overwhelming high frequency and amplitude noise. If more than 3-s of a channel were marked as artifact, the entire channel's recording was omitted. Channels were montaged using bipolar re-referencing such that the differences between adjacent contacts were taken (e.g. electrode contacts LA01, LA02, LA03, LA04 to LA01-LA02, LA02-LA03, LA03-LA04). Data were then filtered for powerline noise using a second-order infinite impulse response filter with quality factor 100 at 60 Hz and harmonic frequencies 120 Hz and 180 Hz. Zero phase filtering was applied to prevent phase delay. All data were then downsampled from their original sampling frequency to the lowest factor above 200 Hz (e.g. 1024 Hz was downsampled to 256 Hz and 500 Hz was downsampled to 250 Hz). This protocol ensured consistency and minimized computational cost since downstream features only used signals between 1 Hz and 80 Hz. The mean of each channel in each recording was subtracted from the recording to center each channel around 0 and remove DC offset.

##### Combining HUP and MNI data

Recognizing the limitations of a single-center approach and to best approximate normal brain electrophysiology, we supplemented our data with data available from the MNI Open iEEG Atlas[13,27,28]. This consisted of normal awake and sleep iEEG from four epilepsy centers. From our data and the MNI Open Atlas, we selected up to 5 minutes of data per sleep and awake stage, limited by detection of sleep with the sleep stage classifier. Following the extraction of bandpower and coherence features, we observed medium effect sizes in features between data collected from HUP and the MNI Open iEEG database, suggesting that some multi-center features could be harmonized (**Supplementary Figure 1**).

### Results

| | Custom Regions | DKT Regions |
|---|---|---|
| 0 | Amyg_Hipp_L | left amygdala, left hippocampus |
| 1 | Amyg_Hipp_R | right amygdala, right hippocampus |
| 2 | Cingulum_L | left caudal anterior cingulate, left isthmus cingulate, left posterior cingulate, left rostral anterior cingulate |
| 3 | Cingulum_R | right caudal anterior cingulate, right isthmus cingulate, right posterior cingulate, right rostral anterior cingulate |
| 4 | FMO_Rect_L | left medial orbitofrontal |
| 5 | FMO_Rect_R | right medial orbitofrontal |
| 6 | Frontal_Mid_All_L | left caudal middle frontal, left rostral middle frontal |
| 7 | Frontal_Mid_All_R | right caudal middle frontal, right rostral middle frontal |
| 8 | Frontal_Sup_All_L | left superior frontal |
| 9 | Frontal_Sup_All_R | right superior frontal |
| 10 | Frontal_inf_All_L | left lateral orbitofrontal, left pars opercularis, left pars orbitalis, left pars triangularis |
| 11 | Frontal_inf_All_R | right lateral orbitofrontal, right pars opercularis, right pars orbitalis, right pars triangularis |
| 12 | Fusiform_L | left fusiform |

| | | |
|---|---|---|
| **13** | Fusiform_R | right fusiform |
| **14** | Insula_L | left insula |
| **15** | Insula_R | right insula |
| **16** | Occipital_Lat_L | left lateral occipital |
| **17** | Occipital_Lat_R | right lateral occipital |
| **18** | Occipital_Med_L | left cuneus, left lingual, left pericalcarine |
| **19** | Occipital_Med_R | right cuneus, right lingual, right pericalcarine |
| **20** | ParaHippocampal_L | left entorhinal, left parahippocampal |
| **21** | ParaHippocampal_R | right entorhinal, right parahippocampal |
| **22** | Parietal_Sup_Inf_L | left inferior parietal, left superior parietal |
| **23** | Parietal_Sup_Inf_R | right inferior parietal, right superior parietal |
| **24** | Postcentral_L | left postcentral |
| **25** | Postcentral_R | right postcentral |
| **26** | Precentral_L | left precentral |
| **27** | Precentral_R | right precentral |
| **28** | Precuneus_PCL_L | left paracentral, left precuneus |
| **29** | Precuneus_PCL_R | right paracentral, right precuneus |
| **30** | SupraMarginal_Angular_L | left supramarginal |
| **31** | SupraMarginal_Angular_R | right supramarginal |
| **32** | Temporal_Inf_L | left inferior temporal |
| **33** | Temporal_Inf_R | right inferior temporal |
| **34** | Temporal_Mid_L | left middle temporal |
| **35** | Temporal_Mid_R | right middle temporal |

| | | |
|---|---|---|
| **36** | Temporal_Sup_L | left superior temporal, left transverse temporal |
| **37** | Temporal_Sup_R | right superior temporal, right transverse temporal |
| **38** | thalam_limbic_L | left accumbens area, left caudate, left pallidium, left putamen, left thalamus proper |
| **39** | thalam_limbic_R | right accumbens area, right caudate, right palladium, right putamen, right thalamus proper |

**Table S1.**

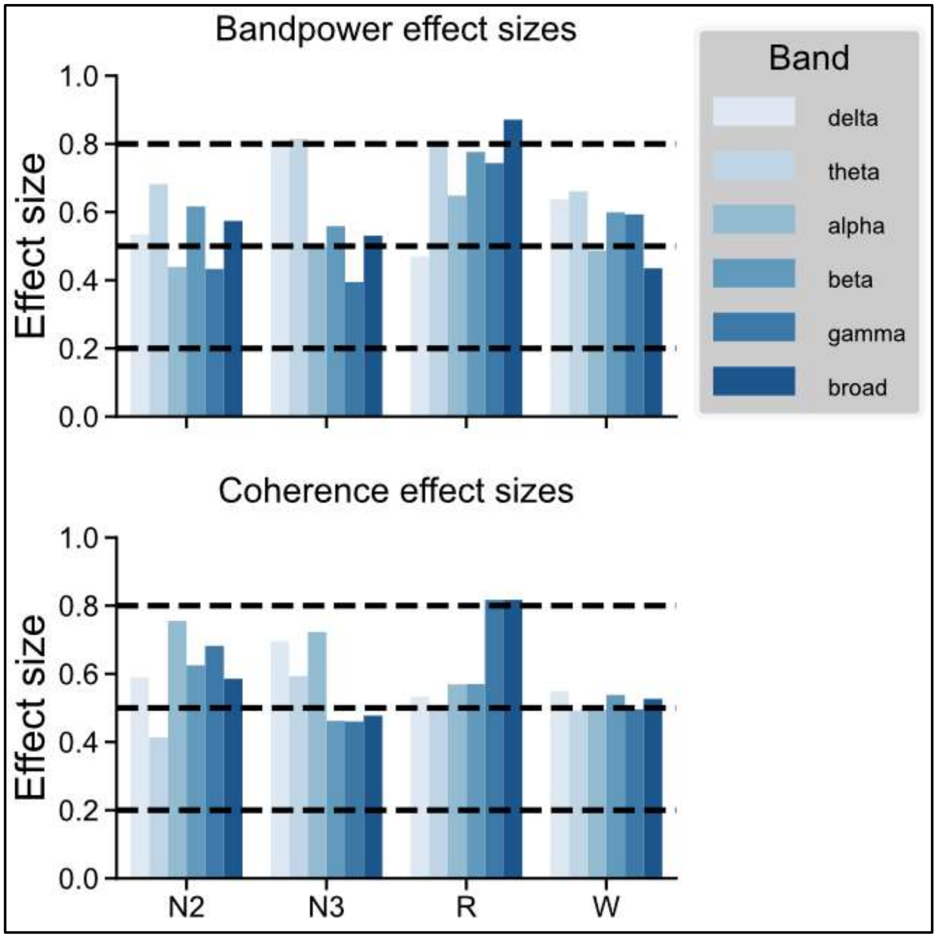


**Figure S1. Effect sizes between centers.** We computed the effect sizes of bandpower and coherence features between normal channels in the HUP and MNI cohorts. Cohen's $d$ for small (0.2), medium (0.5) and large (0.8) effects are shown.

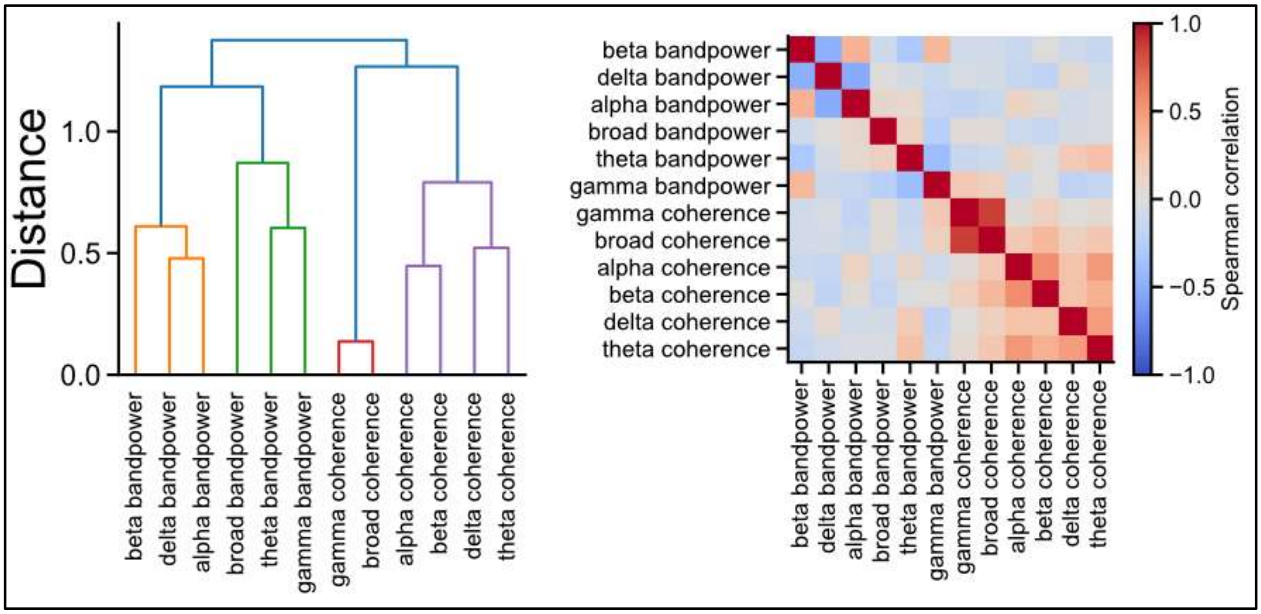


**Figure S2. Similarity of univariate and bivariate features.** Left) Hierarchical clustering of *z*-scores revealed clustered features with bandpower and coherence clusters forming the first link. Right) Further clustering and visualization of pairwise Spearman correlations revealed low and high frequency coherence clusters.

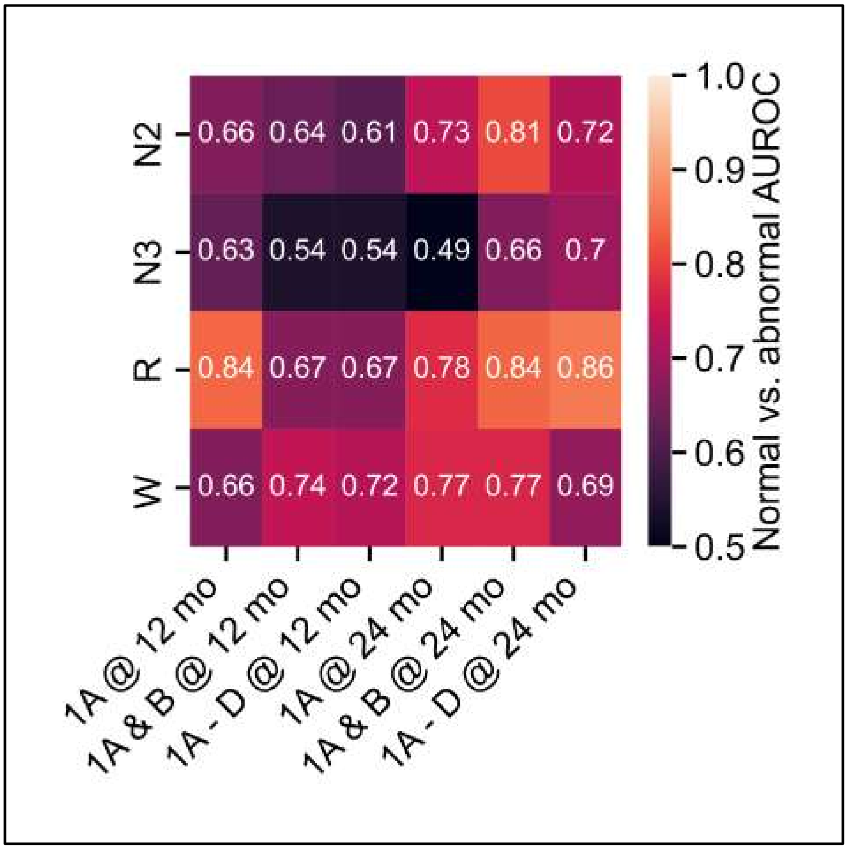

**Figure S3. Predicting abnormalities across outcome thresholds.** Predicting abnormalities at the channel level requires fixing a threshold for good vs. poor surgical outcome of seizure freedom. We applied a parameter sweep of where we separated good and poor outcomes and re-trained the random forest model on the *z*-score features for each channel. All models where good outcome is set as Engel 1 at 24 months had comparably strong performance.

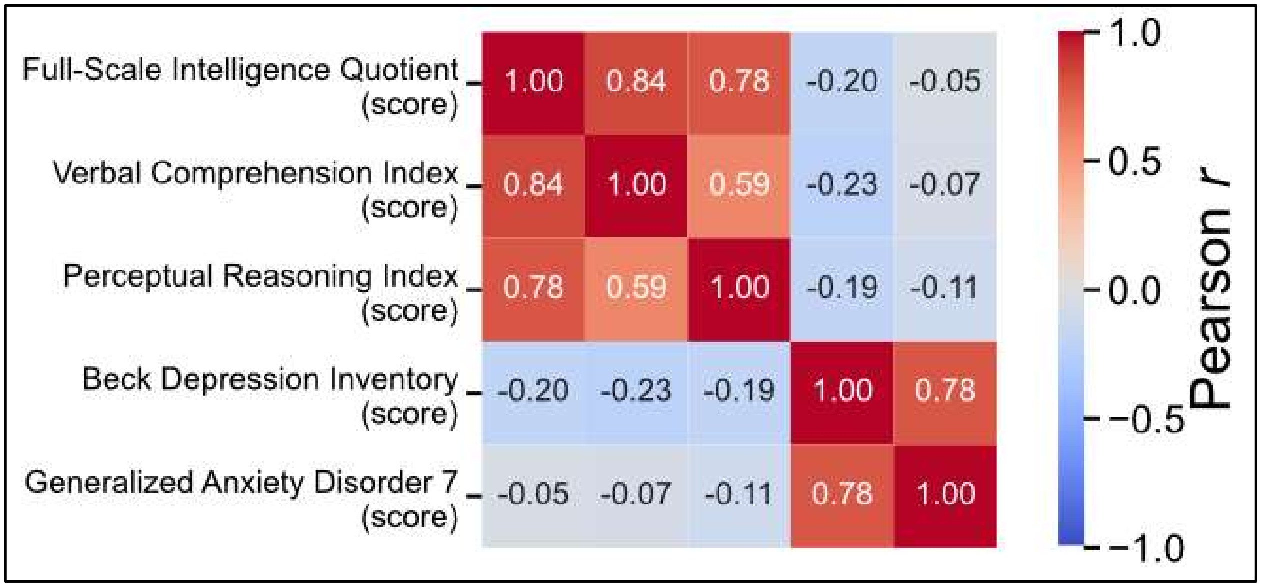


**Figure S4. Clustering of neuropsychological test performance.** To minimize the number of necessary comparisons while adequately representing the diversity of neuropsychological exams, we competed pairwise Pearson correlations between the 5 reported exams. We observed two clusters: Cluster 1 consisted of the Full-Scale Intelligence Quotient, Verbal Comprehension Index, and Perceptual Reasoning Index; Cluster 2 consisted of the Beck Depression Inventory and Generalized Anxiety Disorder.

| | Therapy | Implant | Sex | Target | Laterality | MRI Lesion Status | Age | Engel Score (12 mo) | Engel Score (24 mo) | Good outcome | Has N2 | Has N3 | Has R | Has W |
|---|---|---|---|---|---|---|---|---|---|---|---|---|---|---|
| 0 | Resection | ECoG | Female | Temporal | L | Non-Lesional | 20-29 | 2.1 | 2.1 | FALSE | TRUE | TRUE | TRUE | TRUE |
| 1 | Resection | ECoG | Male | Frontal | L | Lesional | 20-29 | 1.1 | 1.4 | TRUE | TRUE | TRUE | TRUE | TRUE |
| 2 | Resection | ECoG | Female | Temporal | R | Lesional | 20-29 | 1.2 | 1.2 | TRUE | TRUE | FALSE | FALSE | TRUE |
| 3 | Resection | ECoG | Female | Temporal | L | Non-Lesional | 30-39 | 1.1 | 1.1 | TRUE | TRUE | TRUE | FALSE | TRUE |
| 4 | Resection | ECoG | Male | Temporal | R | Non-Lesional | 20-29 | 1.2 | 1.2 | TRUE | TRUE | TRUE | TRUE | TRUE |
| 5 | Resection | ECoG | Male | Frontal | R | Non-Lesional | 30-39 | 1.4 | 1.3 | TRUE | TRUE | FALSE | FALSE | TRUE |
| 6 | Resection | ECoG | Female | Temporal | L | Lesional | 20-29 | 1.1 | 1.1 | TRUE | TRUE | TRUE | FALSE | TRUE |
| 7 | Resection | SEEG | Male | Other | R | Lesional | 30-39 | 1.1 | 1.1 | TRUE | TRUE | TRUE | FALSE | TRUE |
| 8 | Resection | ECoG | Male | Temporal | R | Non-Lesional | 30-39 | 1.1 | 1.1 | TRUE | TRUE | TRUE | TRUE | TRUE |
| 9 | Resection | ECoG | Female | Temporal | L | Non-Lesional | 40-49 | 1.2 | 1.2 | TRUE | FALSE | TRUE | FALSE | TRUE |
| 10 | Resection | ECoG | Female | Temporal | L | Non-Lesional | 20-29 | 3.1 | 2.3 | FALSE | TRUE | TRUE | TRUE | TRUE |
| 11 | Resection | SEEG | Male | Other | R | Lesional | 20-29 | 1.1 | 1.1 | TRUE | TRUE | TRUE | TRUE | TRUE |

| | | | | | | | | | | | | | | |
|---|---|---|---|---|---|---|---|---|---|---|---|---|---|---|
| 12 | Ablation | SEEG | Female | Temporal | R | Lesional | 50-59 | 1.1 | 1.1 | TRUE | TRUE | TRUE | FALSE | TRUE |
| 13 | Resection | ECoG | Female | Temporal | R | Non-Lesional | 40-49 | 1.1 | 1.1 | TRUE | TRUE | TRUE | TRUE | TRUE |
| 14 | Ablation | SEEG | Male | Temporal | R | Non-Lesional | 30-39 | 2.1 | 2.2 | FALSE | TRUE | TRUE | TRUE | TRUE |
| 15 | Ablation | SEEG | Male | Temporal | L | Lesional | 30-39 | 2.1 | 4.3 | FALSE | TRUE | TRUE | FALSE | TRUE |
| 16 | Ablation | SEEG | Male | Temporal | R | Non-Lesional | 30-39 | 2.3 | 1.3 | FALSE | TRUE | TRUE | FALSE | TRUE |
| 17 | Ablation | SEEG | Male | Temporal | L | Lesional | 30-39 | 1.2 | 1.4 | TRUE | TRUE | TRUE | FALSE | TRUE |
| 18 | Resection | SEEG | Male | Temporal | R | Non-Lesional | 19-Oct | 1.1 | 1.1 | TRUE | TRUE | TRUE | TRUE | TRUE |
| 19 | Ablation | SEEG | Male | Frontal | R | Non-Lesional | 30-39 | 1.1 | 2.1 | FALSE | TRUE | FALSE | TRUE | TRUE |
| 20 | Ablation | SEEG | Female | Temporal | R | Non-Lesional | 20-29 | 2.2 | 3.1 | FALSE | TRUE | TRUE | FALSE | TRUE |
| 21 | Ablation | SEEG | Male | Frontal | L | Non-Lesional | 40-49 | 3.1 | 3.1 | FALSE | TRUE | TRUE | FALSE | TRUE |
| 22 | Resection | SEEG | Female | Temporal | R | Non-Lesional | 40-49 | 1.1 | 1.1 | TRUE | TRUE | TRUE | TRUE | TRUE |
| 23 | Resection | SEEG | Male | Temporal | L | Non-Lesional | 20-29 | 1.1 | 4.1 | FALSE | TRUE | TRUE | TRUE | TRUE |
| 24 | Resection | SEEG | Female | Temporal | R | Non-Lesional | 40-49 | 1.1 | 1.4 | TRUE | TRUE | TRUE | TRUE | TRUE |
| 25 | Ablation | SEEG | Female | Frontal | L | Non-Lesional | 20-29 | 1.2 | 2.1 | FALSE | TRUE | TRUE | TRUE | TRUE |

| | | | | | | | | | | | | | | |
|---|---|---|---|---|---|---|---|---|---|---|---|---|---|---|
| 26 | Ablation | SEEG | Female | Temporal | L | Lesional | 30-39 | 2.2 | 2.2 | FALSE | TRUE | TRUE | TRUE | TRUE |
| 27 | Ablation | SEEG | Male | Temporal | L | Non-Lesional | 20-29 | 1.2 | 1.2 | TRUE | TRUE | TRUE | TRUE | TRUE |
| 28 | Ablation | SEEG | Male | Temporal | R | Non-Lesional | 20-29 | 1.2 | 1.1 | TRUE | TRUE | TRUE | TRUE | TRUE |
| 29 | Ablation | SEEG | Male | Temporal | L | Non-Lesional | 20-29 | 4.1 | 4.1 | FALSE | TRUE | TRUE | TRUE | TRUE |
